# North-south pathways, emerging variants, and high climate suitability characterize the recent spread of dengue virus serotypes 2 and 3 in the Dominican Republic

**DOI:** 10.1101/2024.02.14.24302795

**Authors:** Isaac Miguel, Edwin P. Feliz, Robinson Agramonte, Pedro V. Martinez, Carlos Vergara, Yvonne Imbert, Lucia De la Cruz, Nurys de Castro, Odalis Cedano, Yamilka De la Paz, Vagner Fonseca, Gilberto A. Santiago, Jorge L. Muñoz-Jordán, Armando Peguero, Robert Paulino-Ramírez, Nathan D. Grubaugh, Ana Maria Bispo de Filippis, Luiz Carlos Junior Alcantara, Jairo Mendez Rico, José Lourenço, Leticia Franco, Marta Giovanetti

## Abstract

We employ a multidisciplinary approach, integrating genomics and epidemiology, to uncover recent dengue virus transmission dynamics in the Dominican Republic. Our results highlight a previously unknown north-south transmission pathway within the country, with the co-circulation of multiple virus lineages. Additionally, we examine the historical climate data, revealing long-term trends towards higher theoretical potential for dengue transmission due to rising temperatures. These findings provide information for targeted interventions and resource allocation, informing as well towards preparedness strategies for public health agencies in mitigating climate and geo-related dengue risks.

## Main text

Dengue fever, caused by dengue virus serotypes 1-4 (DENV-1-4), represents a significant public health challenge in tropical and subtropical regions (1). A vector-borne disease predominantly transmitted by *Aedes aegypti* mosquitoes, annually affects millions with symptoms ranging from mild fever to severe conditions like dengue hemorrhagic fever and dengue shock syndrome (1). In the past 3 years, dengue activity has surged in the Americas. Approximately 3.4 million cases were reported in 2023, exceeding the 2.8 million total for 2022 (2). Situated in the Caribbean basin, the Dominican Republic has been frequently grappled with dengue outbreaks over the past two decades. The high incidence of dengue in this country is thought to impact the overall epidemiology of dengue in the region.

In 2023, 28,078 cases have been reported by the country’s surveillance system which captures physicians’ reporting of dengue cases (2). Though this surveillance system does not provide a complete picture of dengue transmission, it allows health authorities to maintain awareness of, and response to, dengue transmission throughout the country. In fact, the number of RT-PCR positive samples normally obtained per year, and exceeding 7,000 in 2023, is sufficiently high to provide an opportunity to further characterize transmission using genomic surveillance approaches or through additional analysis of related epidemiological data. In this regard, we implemented a genome-based surveillance approach in partnership with the Pan-American Health Organization and the Central Public Health Laboratory in Santo Domingo to sequence and analyze DENV whole genome sequences from the 2023 epidemic. We aimed and identifying existing lineages within the context of recent emergence of novel DENV-2 and -3 variants in the Americas (3). In addition, by integrating genomic, epidemiological, and climatic data, we provide a historical overview of the epidemiological trajectory of DENV in the Dominican Republic. This comprehensive analysis lays the groundwork for establishing a genomic surveillance model in the Caribbean, with potential to monitor and respond to dengue transmission in the region.

## Results

A total of 85 complete genomes were obtained from 100 PCR positive samples with sufficient DNA (2 ng/L) for library preparation. The average cycle threshold (Ct) value for PCR was 24.6, with values ranging between 12.6 and 36 (**Table S1**). The ages of the patients sampled ranged from 2 to 48 years, with a median age of 17 years. Of these patients, 53% (n=45) were male (**Table S1**), and all cases examined were classified as autochthonous. The sequencing procedure yielded an average coverage of 84%, ranging from 60% to 99% (**Table S1**). This allowed for the identification of the DENV-2 genotype III variant (n=29) and the DENV-3 genotype III (n=56) variant. Genome sequences were obtained from all three macro-regions of the Dominican Republic, which are further segmented into ten specific regions (**Figure 1A**).

**Figure 1.**
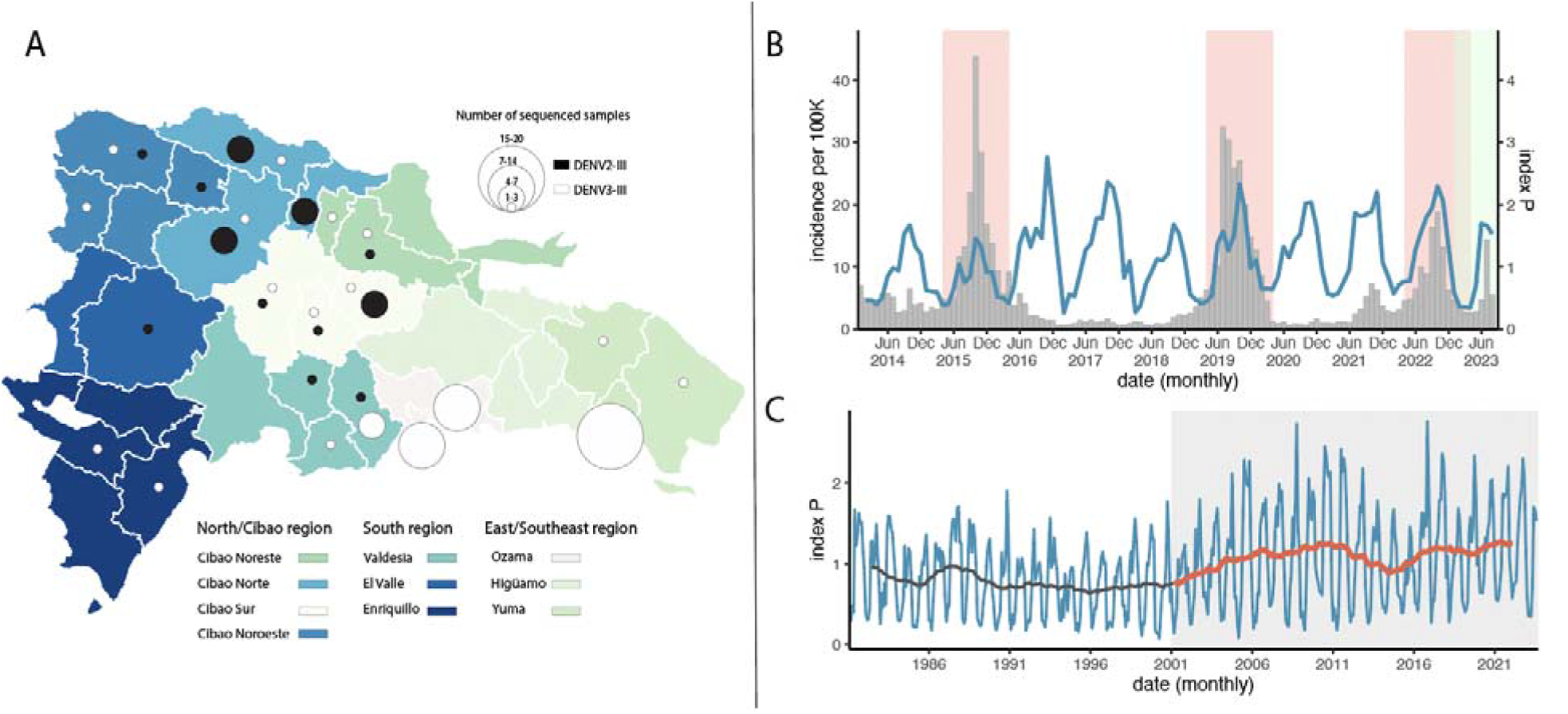
Dynamics of DENV-2 and DENV-3 in the Dominican Republic. A) Map of the Dominican Republic (DR) depicting the sampling of new DENV-2 and DENV-3 genome sequences by region and district. The color and size of the circles represent the number of new genomes generated in this study: black for DENV-2 and white for DENV-3. B) Time series of dengue cases reported monthly in Dominican Republic, presented as incidence per 100,000 inhabitants (gray bars) versus the climate-based suitability (index P, blue line) for the period data where available. The shaded areas in pink mark visually identifiable waves of cases (between April of consecutive years, see main text) and green marks the year 2023 from which all genomic samples were obtained. C) Long-term time series of monthly climate-based suitability (index P, 1981-2023, blue line). The moving average of suitability (plus-minus 18 months) is presented in black before the year 2001 and in red thereafter (the shaded gray area highlights this time period separation).

The distribution of the DENV-2-III and DENV3-III genotypes was geographically distinct. DENV-2-III was predominant in the north-central parts of the country. In contrast, DENV-3-III was found in the north-central and southeastern regions (**Figure 1A**).

While climate-based suitability for DENV transmission presented clear, yearly oscillations demarking time periods with potential for dengue activity, notified DENV cases (between 2014 and 2023) revealed only three waves, of cases (defined as increases in the number of cases above the historical median) occurring in 2015-2016, 2019-2020 and 2021-2022 (**Figure 1B**). The reasons why transmission is low in some years are unclear, though we can consider a mix of local factors including accumulated herd-immunity to specific serotypes, yearly changes in mosquito populations not captured by the climate-based suitability measure, and population cross-immunity post Zika virus emergence after 2016 (4-7). Nonetheless, during the three epidemic waves, Pearson’s correlation between cases and suitability was high, at 0.88 for 2015-2016, 0.76 for 2019-2020 and 0.93 for 2021-2022. Together, these results suggest that, although climate suitability alone is insufficient to promote waves of DENV transmission, local temperature variation tends to be highly associated.

We also explored the historical trends in climate-based suitability in the Dominican Republic (over the past 40 years, **Figure 1C**). This study revealed recent trends towards higher suitability for DENV transmission, particularly after the turn of the century, which derives from the emergence of much higher seasonal peaks of suitability; before 2001, the average yearly peak of suitability was 1.42 (min 0.95; max 1.89), while after 2001, the average peak was 2.08 (min 1.45; max 2.76). However, the time period analyzed is too short for a statistical assessment of significance. Local climate change, driven by rises in temperature, is theoretically favoring the transmission of DENV in the Dominican Republic, as in other countries of the region.

To understand the phylogenetic history of DENV-2-III, we combined our recently sequenced samples (n=23) from 2023 with an additional set of 14 DENV-2-III viral genomes that were sequenced in the Dominican Republic in 2022 (8). This set included samples collected from the Southern, Eastern regions, and the National District (Santo Domingo) (8). These sequences were then combined with other sequences of the same genotype (n=647) retrieved from GenBank for context. Our analysis revealed that the new genomes from this study cluster with sequences recently obtained by others (8) and form a distinct monophyletic clade with robust statistical support that is basal to the DENV-2-III BR4 variant that emerged in 2019 in Brazil (**Figure 2**) (9).

**Figure 2.**
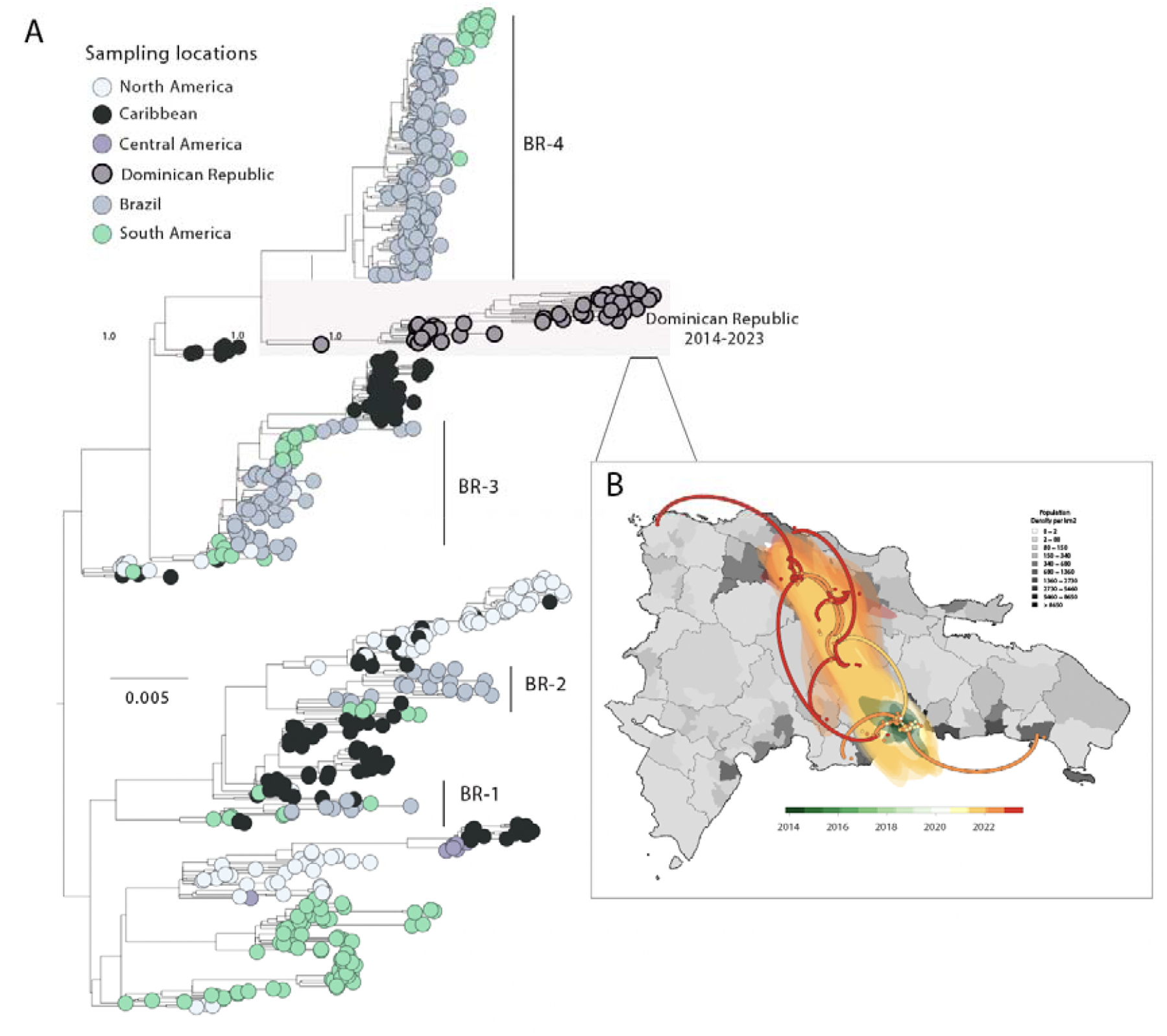
Dispersion dynamics of DENV-2-III in the Dominican Republic. **A)** Maximum likelihood (ML) phylogenetic analysis of 29 new complete genome sequences of DENV-2-III generated in this study combined with 661 sequences from GenBank. The scale bar represents units of nucleotide substitutions per site (s/s) and the tree is mid-pointed rooted. Colors represent geographic sampling locations. B) The highlight on the right (panel B) shows the phylogeographic reconstruction of the Dominican Republic Clade (n=43). Solid curved lines denote the links between nodes and the directionality of movement. Circles represent nodes of the MCC phylogeny and are colored according to their inferred time of occurrence. Shaded areas represent the 80% highest posterior density interval and depict the uncertainty of the phylogeographic estimates for each node.

Additionally, we reconstructed the viral movements of this clade (n=43) within the different regions in the Dominican Republic. We estimate that the mean time of origin of this variant was early-January 2014 with a 95% highest posterior density (HPD), ranging from mid-June 2013 to late-January 2014. These results suggest that the 2023 epidemic may have not been caused by a novel introduction but could be the result of continual transmission within the Dominican Republic of a viral strain that was introduced in the region in early 2014. This variant spread from the southern part of the country (Ozama region) toward the southeastern, northern and midwestern, as demonstrated in **Figure 2**.

To explore the phylogenetic history of DENV-3-III in the Dominican Republic, we combined our new genome dataset with 1,760 sequences from GenBank for global context. Our analysis revealed that the new genomes group within the recently reported American lineage II of genotype III; however, our sequences formed a distinct monophyletic clade that separates from the main lineage, which includes viral sequences recently identified from other Caribbean and Latin American countries, such as Cuba, Puerto Rico, Brazil, Suriname, and as well as in North America (specifically Florida) (3, 8). These results propose a complex transmission scenario, with introduction events likely mediated by trans-continental travel and underscores the significant influence of human movement in facilitating the spread and introduction of viral lineages to new regions (8) (**Figure 3**).

**Figure 3.**
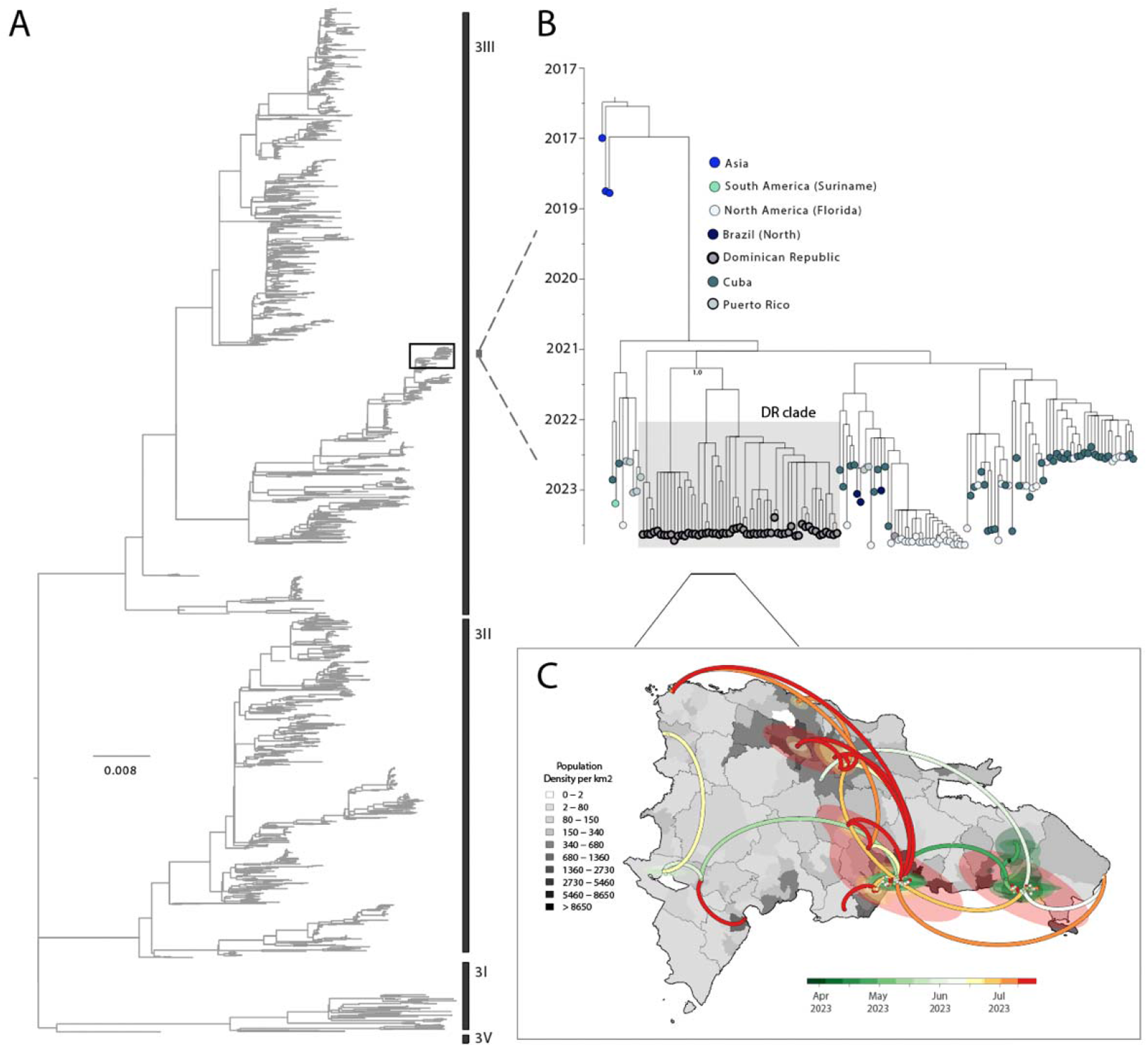
Dispersion dynamics of DENV-3-III in the Dominican Republic. **A)** Maximum Likelihood (ML) phylogenetic analysis of 56 new complete genome sequences of the DENV-3-III genotype from this study, in addition with 1,760 sequences from GenBank representing various DENV-3 genotypes. The scale bar represents units of nucleotide substitutions per site (s/s) and the tree is mid-pointed rooted; B) Maximum clade credibility tree (MCC) of the DENV-3-III emerging American lineage II from 2022-2023, including n=56 complete genome sequences from the Dominican Republic from this study combined with 168 additional genomes for context. Sequences are colored according to sampling location. C) Phylogeographic reconstruction of the Dominican Republic Clade (n=56). Circles represent nodes of the MCC phylogeny and are colored according to their inferred time of occurrence. Shaded areas represent the 80% highest posterior density interval and depict the uncertainty of the phylogeographic estimates for each node. Solid curved lines denote the links between nodes and the directionality of movement.

We investigated the spatial-temporal dynamics of the Dominican Republic DENV-3-III variant in more detail using a smaller data set (n=56) derived from the American lineage II. Phylogeographic analyses allowed the reconstruction of viral movements across different regions in the Dominican Republic (**Figure 3**) and suggested a mean time of origin in early April 2023 (95% highest posterior density (HPD): 2 to 27 April 2023). This variant spread from the south of the country (Yuma region) towards the north and later to the southern part of the country, as indicated by isolates from the Cibao Noroeste, El Valle, and Enriquillo regions (**Figure 3**).

## Conclusion

In conclusion, the implementation of genomic surveillance and its integration into the national laboratory surveillance of dengue produced valuable results that represent a significant step forward in our understanding of the recent epidemiology of the dengue virus in the Dominican Republic. This study revealed the likely-persistence of a variant of DENV-2-III, the recent emergence of variant of DENV-3-III American lineage II and produced an unprecedented number of complete genomes from the Dominican Republic, which will contribute to future research in the country and the Caribbean. We have also unveiled the co-circulation of different DENV serotypes and genotypes, identified a transmission pathway that appears to run along the North-South axis of the country, investigated the impact of climate on the transmission potential of DENV. The North and South of the country could be considered points of entry of viral variants due to the presence of international airports and cruise ports. Our study identified the South of the country as the potential entry point for the DENV-2-III and DENV-3-III variants, thus entry and viral spread from these points merits further study.

A notable discovery of our study is the geographically distinct distribution of the DENV-2-III and DENV-3-III genotypes. DENV-2-III primarily prevails in the north-central regions, while DENV-3-III is prevalent in both the north-central and southeastern areas. This distribution pattern may be attributed to differing herd immunity landscapes for the two serotypes, variations in transmission dynamics, or viral competition for hosts. This highlights the critical need for a deeper understanding of the local epidemiology of dengue to develop and implement optimized, region-specific intervention strategies.

Seroprevalence studies are also needed to characterize the local immune landscape against the different serotypes and to better understand the spatial distribution of serotypes in the island. This information would prove useful to perceive current impact on regional public health and potential effects of future serotype or genotype switching events. Active tracking of variants, viral molecular evolution, and spread can enable effective and timely public health interventions, ensuring that communities are strengthened against the persistent threat of dengue and fostering adaptability in response to the changing landscape of infectious diseases.

## Material and Methods

### Ethics statement

The Pan American Health Organization Ethics Review Committee (PAHOERC) reviewed and approved this project (Ref. No. PAHO-2016-08-0029). The samples used in this study were de-identified residual samples from the routine diagnosis of arboviruses at the Dominican Republic public health laboratory, which is part of the Dominican Republic Ministry of Health’s public network.

### Sample collection and whole genome sequencing

A total of 85 samples, were retrieved from patients presenting clinical symptoms consistent with dengue viral infection. These samples were sent to the Laboratorio Central de Salud Pública of the Dominican Republic, in Santo Domingo for genomic sequencing. Samples were initially submitted to nucleic acid extraction using the QIAamp Viral RNA Mini Kit (Qiagen). Subsequently, the samples underwent real-time reverse transcription PCR (RT-qPCR) for DENV 1-4, as previously described (9). Positive samples (n=85) displaying a cycle threshold (Ct) value of ≤36, were subjected to a whole genome amplification process using a set of tiled primers previously described (10). The resulting DNA amplicons were purified using AMPure XP beads (Beckman Coulter, Brea, USA). Library preparation of the purified amplicons was conducted using the COVIDseq Kit (Illumina, San Diego, USA) which was originally designed for SARS-CoV-2 genomic research but has been adapted for other viruses (11, 12, 13). The subsequent sequencing process was carried out utilizing the Illumina MiSeq platform (Illumina, San Diego, USA), adhering closely to the manufacturer’s recommended protocols. The generation of consensus sequences was performed using the ViralUnity pipeline (available at https://github.com/filiperomero2/ViralUnity), and the typing assignment of dengue virus sequences was performed using the dengue virus typing tool available at https://www.genomedetective.com/app/typingtool/dengue/ (14).

### Phylogenetic and phylodynamic inferences

We constructed phylogenetic trees to explore the relationship of the sequenced genomes from the Dominican Republic to those of other regions sampled globally. All sequences were aligned using MAFFT (15) and manually edited using AliView (16). Preliminary maximum likelihood phylogenies were built using IQ-TREE 2 software under the HKY+G4 substitution model (17). We inferred time-scaled trees by using TreeTime (18). The BEAST software (19) was employed to infer time-scaled phylogenetic trees, preceded by the use of TempEst (20) to evaluate the existence of a temporal signal. We utilized a rigorous model selection approach employing both path-sampling (PS) and steppingstone (SS) procedures to determine the optimal molecular clock model for the Bayesian phylogenetic analysis (21). The uncorrelated relaxed molecular clock model was selected for all datasets based on the estimation of marginal likelihoods. The estimation additionally utilized the codon-based SRD06 model of nucleotide substitution and the nonparametric Bayesian Skyline coalescent model. To simulate the geographic spread of the identified 2022-2023 transmission clade, we employed a flexible relaxed random walk diffusion model (22, 23). This model accounts for the variation in dispersal rates across different branches, using a Cauchy distribution and a jitter window site of 0.01 (24, 25).

Each sequence was assigned geographic coordinates of latitude and longitude. The Bayesian phylogenetic inference analyses were conducted in BEAST v1.10.4, with two runs of 50 million Markov Chain Monte Carlo chains (MCMC) each, and samples were taken every 10,000 steps in the chain. Convergence for each run was evaluated in Tracer, ensuring that the effective sample size for all significant model parameters was more than 200. The maximum clade credibility (MCC) trees for each run were summarized using TreeAnnotator after removing the initial 10% as burn-in. Ultimately, we employed the R package ‘seraphim’ version 1.0 (25) to extract and visualize spatiotemporal data included inside the posterior trees.

### Eco-epidemiological modelling

The epidemiological data on weekly notified cases of DENV in the Dominican Republic, from 2013 to 2023, were collected and organized from the PAHO data repository for dengue (26). Notified infections are classified as cases of dengue where a diagnostic test has been conducted and yielded a positive result, according to the PAHO platform. We calculated the theoretical appropriateness of climate-based transmission of the dengue virus using the following mathematical expression of index P, where u stands for humidity and t for temperature:

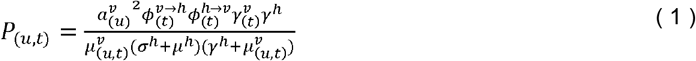

Briefly, the index uses mathematical expressions of empirically demonstrated relationships between DENV and *Ae. aegypti* traits and meteorological variables. Climate-dependent traits include the extrinsic incubation period 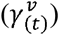, adult mosquito lifespan 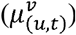, adult mosquito biting rate 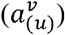, transmission probability per mosquito bite from infected human to susceptible mosquito 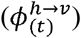 and from infected mosquito to susceptible human 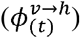. Traits that are climate-independent include intrinsic incubation period (*γ*^*h*^), human lifespan (*μ*^*h*^) and human infectious period (*σ*^*h*^). Full methodological details and technical validation of Index P can be found in Nakase et al (27, 28). Monthly climate data for the Dominican Republic was obtained from Copernicus.eu satellite climate data (29). We derived the temperature data summary by computing the yearly lowest, mean, and maximum values.

## Data Availability

Newly generated sequences have been deposited in GenBank under accession numbers OR616454-OR616538

## Acknowledgement

This study was supported by PAHO, USAID, Dominican Republic Public Health Ministry, (funding for the genomics department of Defillo national Laboratory) and in part by the National Institutes of Health (NIH) USA grant U01 AI151698 for the United World Arbovirus Research Network (UWARN) and the CRP-ICGEB RESEARCH GRANT 2020 Project CRP/BRA20-03, Contract CRP/20/03. M. Giovanetti’s funding is provided by PON “Ricerca e Innovazione” 2014-2020. The authors would also like to acknowledge the Global Consortium to Identify and Control Epidemics – CLIMADE (T.O., L.C.J.A., E.C.H., J.L., M.G.) (https://climade.health/) and National Institute of Allergy and Infectious Diseases of the NIH Award Number DP2AI176740 (NDG). The findings and conclusions in this report are those of the author(s) and do not necessarily represent the official position of the CDC and NIH.

## Author contributions

Conception and design: I.M., J.L., and M.G.; Investigations: I.M., E.F., R.A., P.V.M., C.V., Y.I., L.d.l.C., N.d.C., O.C., Y.D.l.P., V.F., G.A.S., J.L.M.J., R.P.R., N.D.G., A.M.B.d.F., L.C.J.A., J.M.R., J.L., L.F., M.G.; Data Analysis: I.M., E.F., V.F., J.L., and M.G; Visualization: V.F., J.L., and M.G; Writing – Original: J.L., and M.G.; Revision: I.M., E.F., R.A., P.V.M., C.V., Y.I., L.d.l.C., N.d.C., O.C., Y.D.l.P., V.F., G.A.S., J.L.M.J., R.P.R., N.D.G., A.M.B.d.F., L.C.J.A., J.M.R., J.L., L.F., M.G.; Resources: I.M., R.P.R., N.D.G., J.M.R., and L.F.

